# Negative online experiences and reporting rates in youth with mental health conditions

**DOI:** 10.1101/2025.06.23.25330123

**Authors:** Mirelle Kass, Lindsay Alexander, Tatum Connell, Sara Bagheri Hamaneh, Najuma Abdullah, Michelle Freund, Michael P. Milham, Aki Nikolaidis

## Abstract

**Importance:** Negative online experiences (NOE) are highly prevalent and pose significant dangers for youth with mental health and neurodevelopmental conditions.

**Objective:** Identify risk factors of NOE and reporting barriers in youths.

**Design:** Cross-sectional survey study (January to July 2023) and qualitative follow-up study (March to May 2023).

**Setting:** A community-based sample from the Child Mind Institute’s Healthy Brain Network (HBN), a transdiagnostic research initiative in New York.

**Participants:** A total of 1,009 youth aged 9-15 years (mean [SD] age, 11.79 [1.71] years; 585 [58.55%] male) completed a 310-item quantitative survey. The qualitative follow-up study (n=109) included four administrations of a 3-day online bulletin board.

**Exposure(s):** Not applicable.

**Main Outcomes and Measures:** Main outcomes include prevalence and risk factors of NOE and reporting behaviors and barriers. Qualitative analyses provided insights into youths’ mental models and cognitive schemas about different NOE forms.

**Results:** Of the 1,129 enrolled participants, 1,009 participants completed the study (89.37%). Over one-fourth (265 [26.6%]) encountered NOE in the past year, with 182 [68.7%] reporting multiple incidents. However, only 53 [20.0%] reported the incident. Mental health symptoms (*p*<.01) and parenting styles (*p*<.001) increased the risk of NOE. Factor analyses identified three key reporting barriers: Reporting Process, Reporting Policy, and Emotional Barriers. Age, social aptitude, mental health symptoms, and parenting styles predicted the likelihood of experiencing the reporting barriers. Analyses of the qualitative follow-up study noted that youths’ decision-making process regarding reporting considers three areas: degree of malice, perpetrator’s intent, and included NOE forms. Ambiguity in these areas contributed to higher reporting uncertainty.

**Conclusions and Relevance:** Findings highlight a gap between the prevalence of NOE and reporting rates in youths with mental health and neurodevelopmental conditions. Demographic, clinical, and parenting factors were risk factors of experiencing NOE or a reporting barrier, underscoring the need for targeted and multifaceted solutions. Potential solutions to aid in safer online spaces are proposed for policymakers, technology developers, clinicians, and educators.

**Key points:** *Question:* When encountering negative online experiences (NOE), what influences reporting likelihood in youth with mental health conditions?

*Findings:* High prevalence and low reporting rates of NOE were observed. Analyses identified Reporting Process, Reporting Policy, and Emotional as key reporting barriers. Parenting styles, mental health symptoms, and demographics were associated with increased likelihood of experiencing NOE or the identified barriers. The qualitative follow-up study provided insights into youths’ thought processes regarding NOE.

*Meaning:* Our findings document key reporting barriers in children and adolescents with mental health and neurodevelopmental conditions and potential suggestions to aid in the development of safer online spaces.

## Introduction

The internet has become deeply integrated into our daily lives, particularly for youth. While childhood internet use has been associated with improvements in academic achievement^1,2^, learning development^3^ and digital literacy^4,5^, digital spaces also introduce significant risks to online harassment (e.g., cyberstalking, grooming), especially for younger users^6^. One report of 1,000 children and adolescents, aged 9-17, found that 48% experienced at least one negative or potentially harmful online incident in the past that made them feel uncomfortable^7^.

In the past year, one in four youths (aged 9-16) reported having a “negative online experience” (NOE) – ‘any unwanted or uncomfortable negative experience while online’^8^. These incidents have well-documented psychological and physical effects^9^ and exist in many forms, including impersonation^10^, doxing^11^, outing^12^, trickery^12^, exclusion^12^, flaming^12^, cyberstalking^12^, sexual cyberbullying^13^, and denigration^14^; all of which have been linked to poorer mental health outcomes^9^. Victims of NOE report higher rates of depression^9,15–19^, anxiety^9,18,20^, self-harm^15,18^, post-traumatic stress disorder^19^, and suicidal ideation^9,15,16,18,21,22^. While negative long-term effects (e.g., worsening depression) have been noted by longitudinal studies^23^, few have explored variances among different forms of NOE.

Despite the growing awareness of NOE, there is still limited understanding of how children and adolescents respond to these encounters. The most common advice provided to a victim of NOE is to disclose the incident^24^, as sharing the encounter has been shown to reduce the likelihood of future harassment^25^. Nonetheless, rates of reporting NOE in children and adolescents remain quite low^7,26^, with one study noting that over one in four youths who experienced NOE did not seek help from the platform, an adult, or a peer^7^. Potential reporting barriers that have been noted include anonymity concerns^7^, fear of escalation^27^, feelings of embarrassment^7,26^, and perceived consequences or retaliation from reporting^26^. Additionally, youths’ comfort levels with disclosing NOE have been found to vary by the report type, with a preference for peers over parents and caregivers^20,26,28^ and great discomfort with the platform directly^26^.

Some studies have explored how parent-child relationships may influence a youth’s likelihood of experiencing or reporting NOE^19,29^. They propose that more inconsistent and distant parenting may be a risk factor for experiencing NOE^29,30^, while greater positive attachment may be a protective factor against the negative impacts of NOE^19^. Additionally, increased risk for experiencing NOE has been observed in individuals with preexisting mental health symptoms^29^ and poorer health^19^; however, little is known about how at-risk youth experience different motivators or barriers to reporting^15,16^.

The present study explored NOE and reporting behaviors in 1,009 children and adolescents with mental health and neurodevelopmental conditions from the Healthy Brain Network^31^, a large-scale, community research initiative. A qualitative follow-up study (*N*=109) provided deeper insights into youths’ decision-making processes and mental models regarding NOE and reporting.

## Methods

### Participant recruitment

Parents and/or caregivers of eligible participants of the Child Mind Institute’s Healthy Brain Network (HBN)^31^ were contacted via recruitment emails and phone calls. Appropriate informed consent and assent processes were conducted remotely with the parent and/or caregiver and the participant, respectively. During the assent process, participants were also able to opt-in to be considered for the qualitative follow-up study, if they were to be randomly selected to participate. All study data collection occurred between January to July 2023.

Participants were compensated with an electronic gift card up to $185 ($75 for the quantitative survey and $110 for the qualitative follow-up study). Participants were able to withdraw from the study at any time. This study was approved by the Advarra Institutional Review Board (https://www.advarra.com/) and followed the American Association for Public Opinion Research (AAPOR) reporting guidelines^32^.

### Measures

The present study involved data previously collected via HBN, a 310-item quantitative survey, and a 3-day qualitative follow-up study (30 minutes/day).

To develop a deeper profile of the participants, a subset of data from HBN that included demographic, cognitive, behavioral, and clinical evaluations, were incorporated. Clinical diagnoses identified by HBN’s neuropsychological clinicians were categorized into seven HBN Diagnosis Categories (eTable 1).

The quantitative survey, administered via REDCap^33^, incorporated existing survey questions (eTable 2) and project developed questions designed by mental health experts. Data collected included background and demographic information, parenting styles, general online behavior, prevalence of NOE, reporting behaviors and desired features to promote online safety. (eMethods and eTable 2)

A subset of the sample were randomly selected to participate in one of the four moderated online bulletin boards to explore how youth experience and respond to different forms of NOE (*N*=109; 13.6%) (eMethods and eTable 3).

### Statistical analysis

All statistical analyses occurred between October 2023 and February 2025 and were conducted using R^34^ (packages: lme4^35^ and stats^36^). For all analyses, a 2-sided statistical significance cutoff of *P*C<C.05 was applied. Odds ratios were calculated to evaluate how sample characteristics influenced the likelihood of NOE (Table 2). We split our sample into two demographically matched halves (i.e., age, sex, race matched). We conducted an exploratory factor analysis on the first half of the sample and performed a confirmatory factor analysis on the second split sample to evaluate model fit. Parallel analysis revealed three significant factors emerging that summarized barriers to reporting (Figure 1). Multivariate regression models were conducted to explore predictors of barriers to reporting.

**Figure 1.**
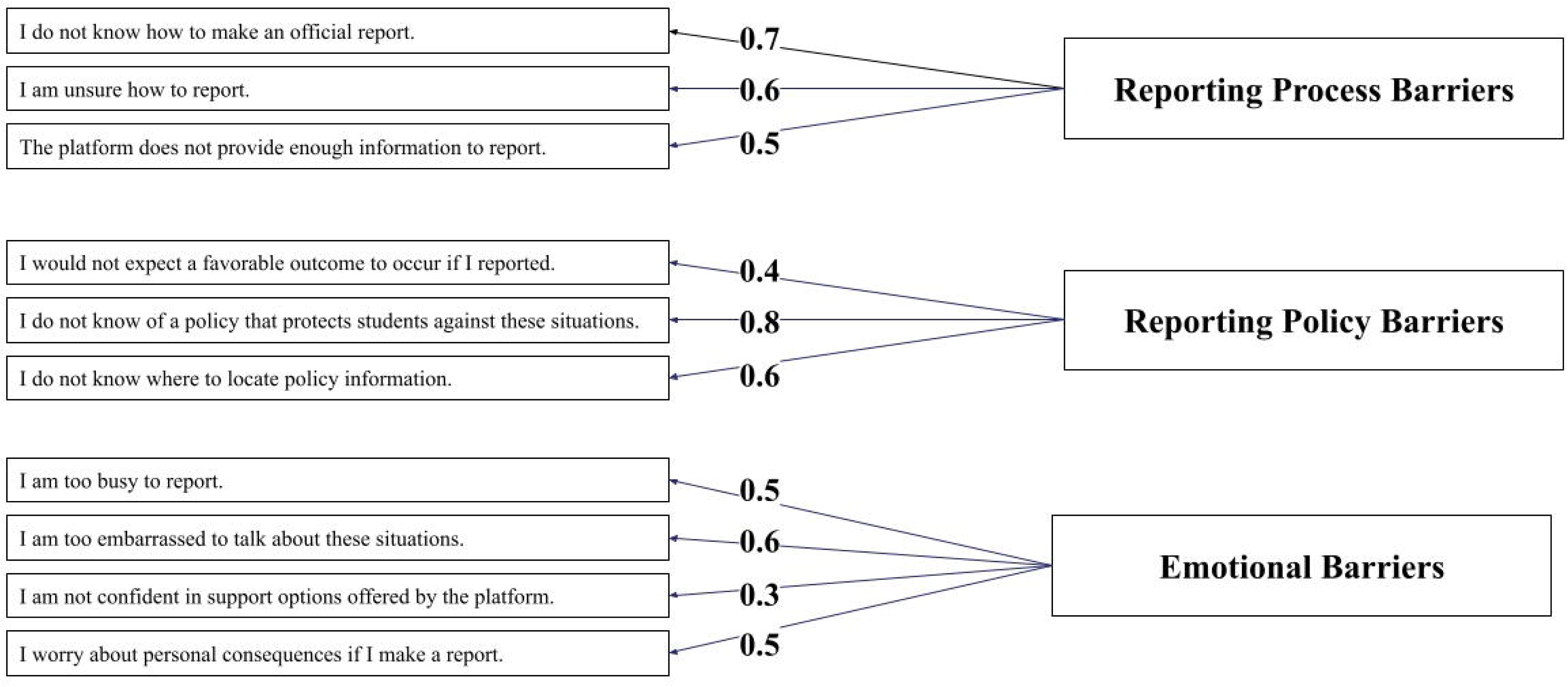
**Factor analysis structure of three reporting barriers.** Exploratory and confirmatory factor analysis results illustrate three distinct barriers to reporting: Process Barriers (difficulty navigating reporting mechanisms), Policy Barriers (uncertainty about a platform’s reporting protocols), and Emotional Barriers (distressing psychological emotions). Standardized factor loadings and model fit indices confirm a robust three-factor structure.

**Table 1.**
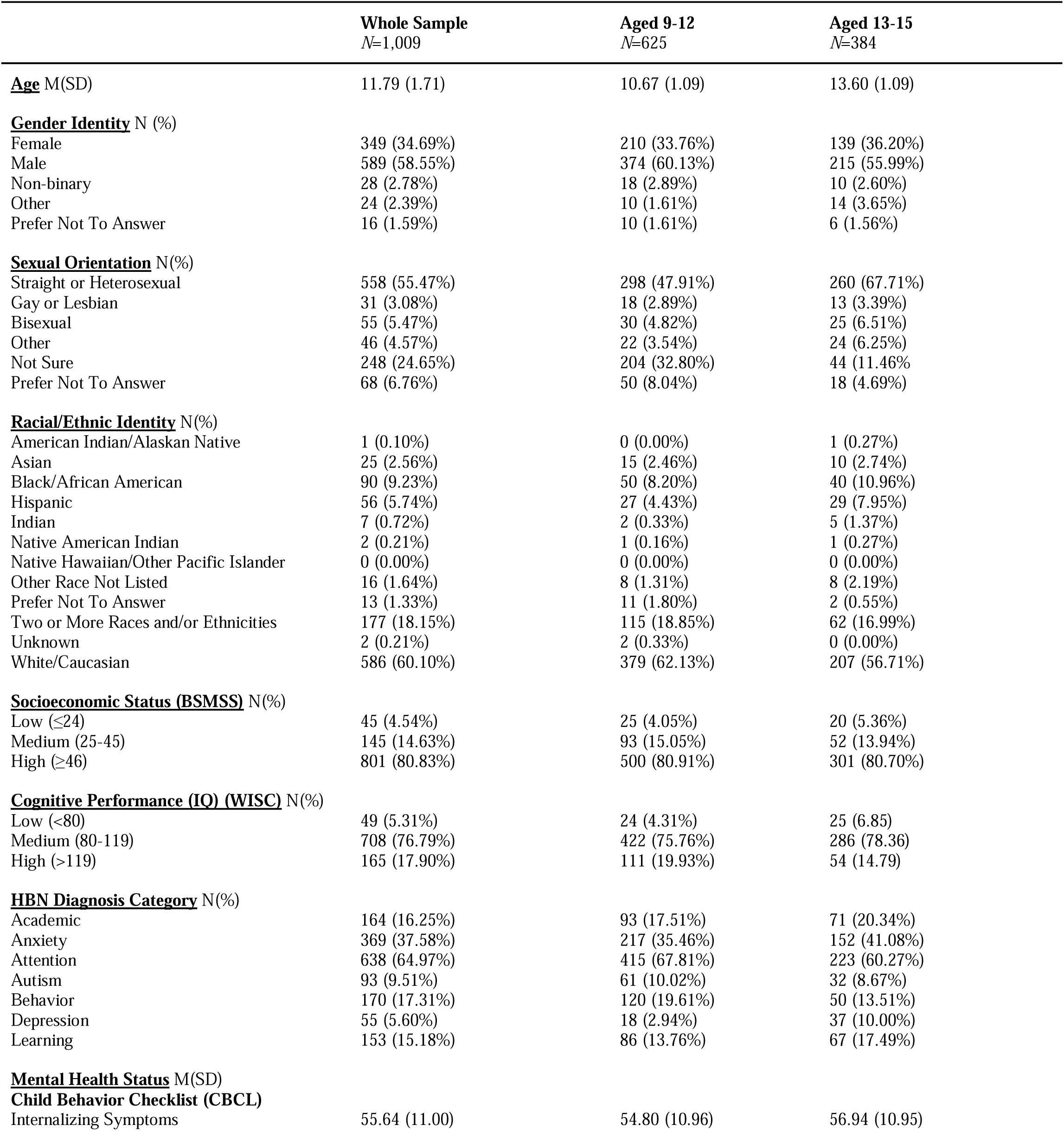
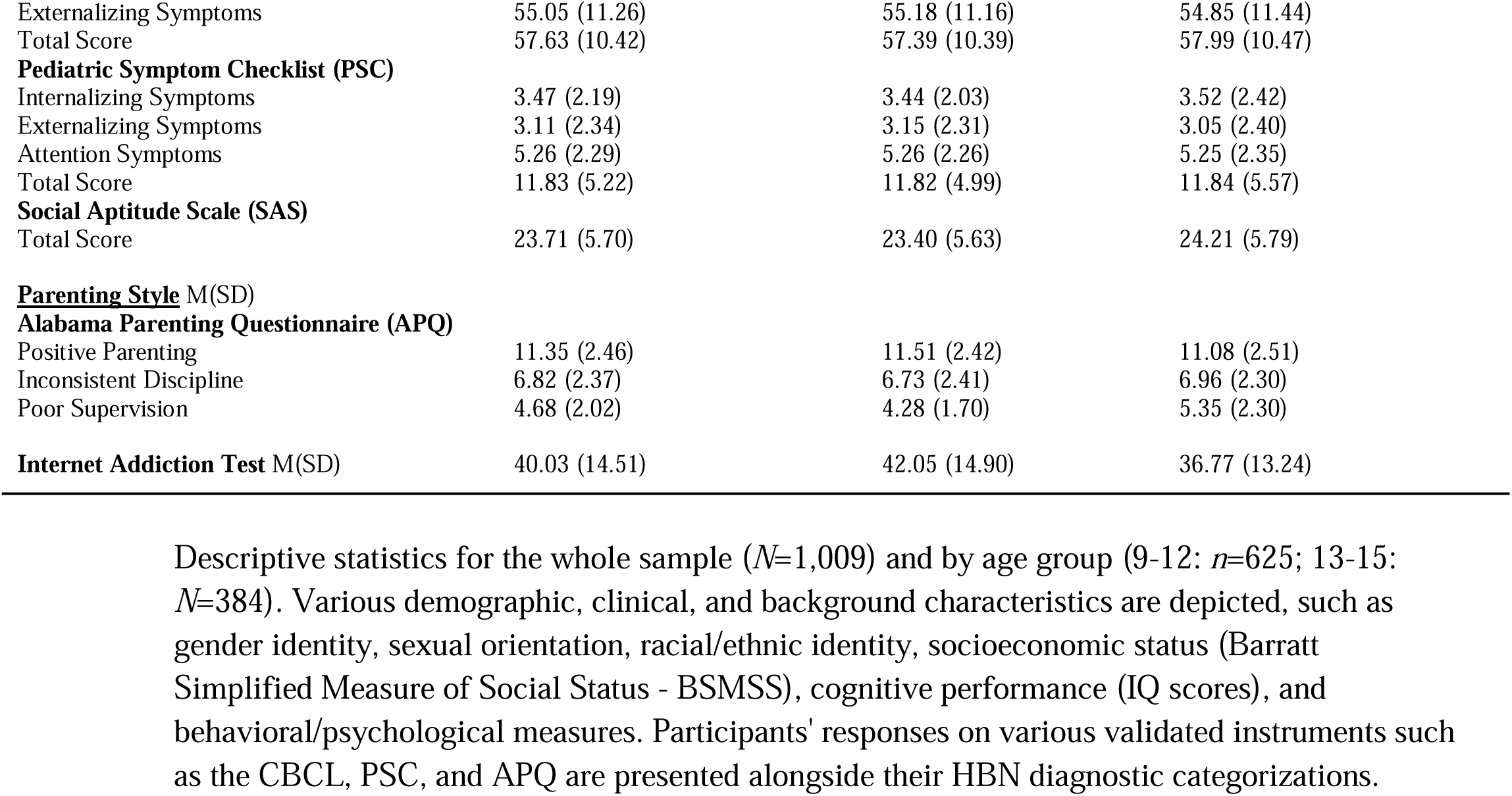
Sample characteristics.

**Table 2.**
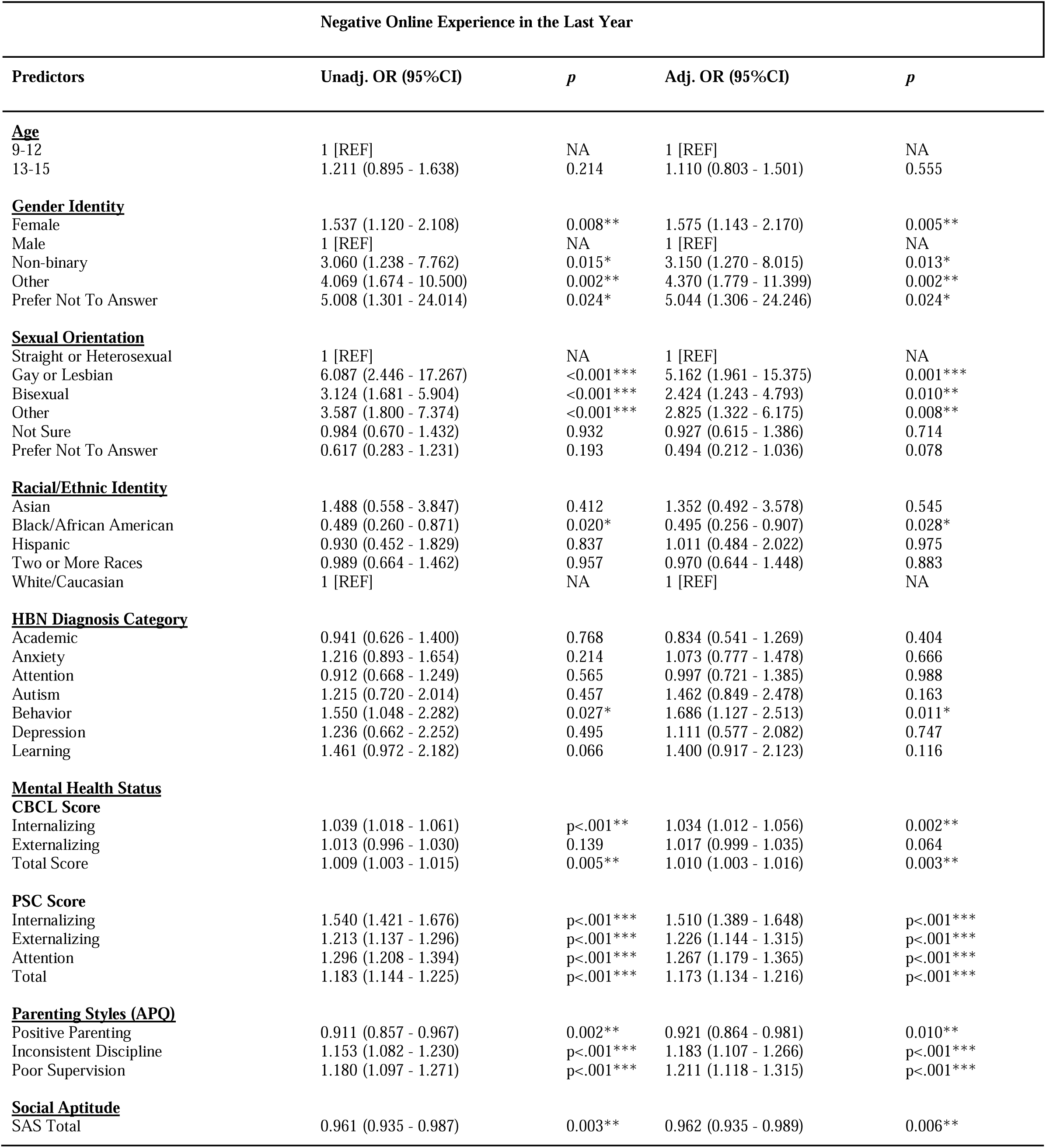
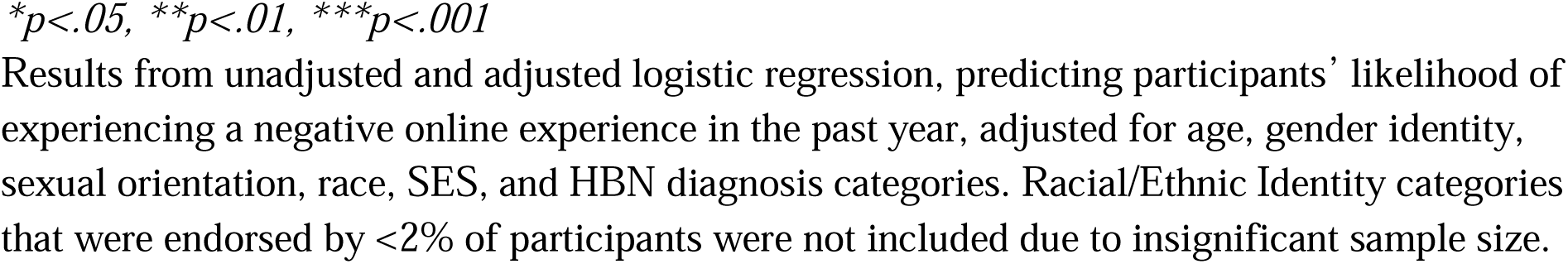
Likelihood of encountering a negative online experience.

| Negative Online Experience in the Last Year |  |  |  |  |
| --- | --- | --- | --- | --- |
| Predictors | Unadj. OR (95%CI) | *p* | Adj. OR (95%CI) | *p* |
| **<u>Age</u>** |  |  |  |  |
| 9-12 | 1 [REF] | NA | 1 [REF] | NA |
| 13-15 | 1.211 (0.895 - 1.638) | 0.214 | 1.110 (0.803 - 1.501) | 0.555 |
| **<u>Gender Identity</u>** |  |  |  |  |
| Female | 1.537 (1.120 - 2.108) | 0.008** | 1.575 (1.143 - 2.170) | 0.005** |
| Male | 1 [REF] | NA | 1 [REF] | NA |
| Non-binary | 3.060 (1.238 - 7.762) | 0.015* | 3.150 (1.270 - 8.015) | 0.013* |
| Other | 4.069 (1.674 - 10.500) | 0.002** | 4.370 (1.779 - 11.399) | 0.002** |
| Prefer Not To Answer | 5.008 (1.301 - 24.014) | 0.024* | 5.044 (1.306 - 24.246) | 0.024* |
| **<u>Sexual Orientation</u>** |  |  |  |  |
| Straight or Heterosexual | 1 [REF] | NA | 1 [REF] | NA |
| Gay or Lesbian | 6.087 (2.446 - 17.267) | <0.001*** | 5.162 (1.961 - 15.375) | 0.001*** |
| Bisexual | 3.124 (1.681 - 5.904) | <0.001*** | 2.424 (1.243 - 4.793) | 0.010** |
| Other | 3.587 (1.800 - 7.374) | <0.001*** | 2.825 (1.322 - 6.175) | 0.008** |
| Not Sure | 0.984 (0.670 - 1.432) | 0.932 | 0.927 (0.615 - 1.386) | 0.714 |
| Prefer Not To Answer | 0.617 (0.283 - 1.231) | 0.193 | 0.494 (0.212 - 1.036) | 0.078 |
| **<u>Racial/Ethnic Identity</u>** |  |  |  |  |
| Asian | 1.488 (0.558 - 3.847) | 0.412 | 1.352 (0.492 - 3.578) | 0.545 |
| Black/African American | 0.489 (0.260 - 0.871) | 0.020* | 0.495 (0.256 - 0.907) | 0.028* |
| Hispanic | 0.930 (0.452 - 1.829) | 0.837 | 1.011 (0.484 - 2.022) | 0.975 |
| Two or More Races | 0.989 (0.664 - 1.462) | 0.957 | 0.970 (0.644 - 1.448) | 0.883 |
| White/Caucasian | 1 [REF] | NA | 1 [REF] | NA |
| **<u>HBN Diagnosis Category</u>** |  |  |  |  |
| Academic | 0.941 (0.626 - 1.400) | 0.768 | 0.834 (0.541 - 1.269) | 0.404 |
| Anxiety | 1.216 (0.893 - 1.654) | 0.214 | 1.073 (0.777 - 1.478) | 0.666 |
| Attention | 0.912 (0.668 - 1.249) | 0.565 | 0.997 (0.721 - 1.385) | 0.988 |
| Autism | 1.215 (0.720 - 2.014) | 0.457 | 1.462 (0.849 - 2.478) | 0.163 |
| Behavior | 1.550 (1.048 - 2.282) | 0.027* | 1.686 (1.127 - 2.513) | 0.011* |
| Depression | 1.236 (0.662 - 2.252) | 0.495 | 1.111 (0.577 - 2.082) | 0.747 |
| Learning | 1.461 (0.972 - 2.182) | 0.066 | 1.400 (0.917 - 2.123) | 0.116 |
| **<u>Mental Health Status</u>** |  |  |  |  |
| **<u>CBCL Score</u>** |  |  |  |  |
| Internalizing | 1.039 (1.018 - 1.061) | p<.001** | 1.034 (1.012 - 1.056) | 0.002** |
| Externalizing | 1.013 (0.996 - 1.030) | 0.139 | 1.017 (0.999 - 1.035) | 0.064 |
| Total Score | 1.009 (1.003 - 1.015) | 0.005** | 1.010 (1.003 - 1.016) | 0.003** |
| **<u>PSC Score</u>** |  |  |  |  |
| Internalizing | 1.540 (1.421 - 1.676) | p<.001*** | 1.510 (1.389 - 1.648) | p<.001*** |
| Externalizing | 1.213 (1.137 - 1.296) | p<.001*** | 1.226 (1.144 - 1.315) | p<.001*** |
| Attention | 1.296 (1.208 - 1.394) | p<.001*** | 1.267 (1.179 - 1.365) | p<.001*** |
| Total | 1.183 (1.144 - 1.225) | p<.001*** | 1.173 (1.134 - 1.216) | p<.001*** |
| **<u>Parenting Styles (APQ)</u>** |  |  |  |  |
| Positive Parenting | 0.911 (0.857 - 0.967) | 0.002** | 0.921 (0.864 - 0.981) | 0.010** |
| Inconsistent Discipline | 1.153 (1.082 - 1.230) | p<.001*** | 1.183 (1.107 - 1.266) | p<.001*** |
| Poor Supervision | 1.180 (1.097 - 1.271) | p<.001*** | 1.211 (1.118 - 1.315) | p<.001*** |
| **<u>Social Aptitude</u>** |  |  |  |  |
| SAS Total | 0.961 (0.935 - 0.987) | 0.003** | 0.962 (0.935 - 0.989) | 0.006** |

*\*p<.05, \*\*p<.01, \*\*\*p<.001*
Results from unadjusted and adjusted logistic regression, predicting participants' likelihood of experiencing a negative online experience in the past year, adjusted for age, gender identity, sexual orientation, race, SES, and HBN diagnosis categories. Racial/Ethnic Identity categories that were endorsed by <2% of participants were not included due to insignificant sample size.

## Results

### Sample characteristics

Of the 2,745 recruited participants, 1,129 enrolled in the study. A total of 135 (11.96%) dropped out or did not complete the survey, consent, or assent. The final study sample included 1,009 participants between the ages of 9 and 15 years old (mean [SD] age, 11.79 [1.71]); 589 (58.6%) were male, 349 (34.7%) were female, 28 (2.8%) were non-binary, 24 (2.4%) listed another gender, and 16 (1.6%) did not report their gender. One participant (0.1%) was American Indian/Alaska Native; 25 (2.6%), Asian; 90 (9.2%), Black/African American; 56 (5.7%), Hispanic; 7 (0.7%), Indian; 2 (0.2%), Native American

Indian; none, Native Hawaiian/Other Pacific Islander; 586 (60.1%), White/Caucasian; 177 (18.2%), Two or More Races and/or Ethnicities; 16 (1.6%) indicated another race or ethnicity not listed; 2 (0.2%) listed unknown, and 13 (1.3%) preferred not to answer. Over half (61.0%) reported household incomes above $100,000. Most participants (898 [90.00%]) satisfied at least one HBN Diagnosis Category; 47.87% (483) satisfied two or more HBN Diagnosis Categories (Table 1).

### General online behavior

Mean total time online per day was 5.94 [4.14] hours. Participants reported spending more time online for personal/social activities (3.60 [2.77] hours) than for school-related activities (2.33 [2.56] hours). Older participants (13-15 years) (7.52 [4.46] hours) reported spending more time online than younger participants (9-12 years) (4.97 [3.60]). Of the online activities assessed, watching online videos (801 [79.85%]) and playing video games (621 [61.85%]) were endorsed most frequently. (eFigure 1)

### Negative online experiences (NOE)

Over one-fourth (265 [26.26%]) reported experiencing at least one NOE in the past year, with 68.68% (182) noting multiple incidents, including on a weekly (11 [4.15%]) or daily (12 [4.53%]) basis. Of the 265 who experienced NOE, most experienced negative emotions during the incident, such as anger (131 [49.43%]), sadness (119 [44.91%]), and fear (115 [43.40%]). Few of those who experienced NOE reported the incident to the platform [20.00%]); most chose to close the platform (98 [36.98%]) or ignore it (79 [29.81%]).

Increased likelihood of NOE was associated with elevated internalizing psychopathology measured by the CBCL (Unadj. OR: 1.03-1.04, 95%CI [1.02, 1.06], *p* <.001; Adj. OR: 1.03, 95%CI [1.01, 1.06], *p*<.01) and the PSC (Unadj. OR: 1.54, 95%CI [1.42, 1.68], *p*<.001; Adj. OR: 1.51, 95%CI [1.39, 1.65], *p*<.001) (Table 2). Higher likelihoods were associated with greater inconsistent discipline and poor supervision scores (Unadj. OR: 1.15-1.18, 95%CI [1.08, 1.27], all *p*<.001; Adj. OR: 1.18-1.21, 95%CI [1.10, 1.32], *p*<.001).

### Barriers to reporting

Parallel analyses identified a three factor solution as the best fit for the reporting barrier questions (Figure 1). Confirmatory factor analysis revealed a good fitting factor structure (CFI≥0.94; TLI≥0.92). The three key reporting barriers were: (1) Reporting Process Barriers; (2) Reporting Policy Barriers; and (3) Emotional Barriers (Figure 1). Two questions loaded separately and were explored independently (eTable 4).

Table 3 shows the multilevel regressions of the three factors. Compared to older participants (13-15 years), younger participants (9-12 years) were significantly more likely to experience all three barriers (Reporting Process Barriers: *b*=0.65, 95%CI [1.02, 0.29], *p*<.001; Reporting Policy Barriers: *b*=0.45, 95%CI [0.81, 0.09], *p*=.013; Emotional Barriers: *b*=0.41, 95%CI [0.75, 0.06], *p*=.020). Lower social aptitude significantly predicted all barriers, but to a lesser extent (Reporting Process Barriers: *b*=0.06, 95%CI [0.09, 0.03], *p*<.001; Reporting Policy Barriers: *b*=0.05, 95%CI [0.08, 0.02], *p*=.001; Emotional Barriers: *b*=0.06, 95%CI [0.08, 0.02], *p*<.001). Participants with elevated internalizing symptoms were more likely to experience Emotional Barriers (*b*=0.09, 95%CI [0.00, 0.17], *p*=.046) only. Positive parenting scores were negatively associated with Reporting Process Barriers (*b*=-0.07, 95%CI [-0.15, 0.00], *p*=.039) and Emotional Barriers (*b*=-0.08, 95%CI [−0.15, −0.01], *p*=.019). Inconsistent discipline scores were positively associated with Reporting Policy Barriers (*b*=0.09, 95%CI [0.01, 0.16], *p*=.026) and Emotional Barriers (*b*=0.08, 95%CI [0.01, 0.16], *p*=.022). (Table 3).

**Table 3.**
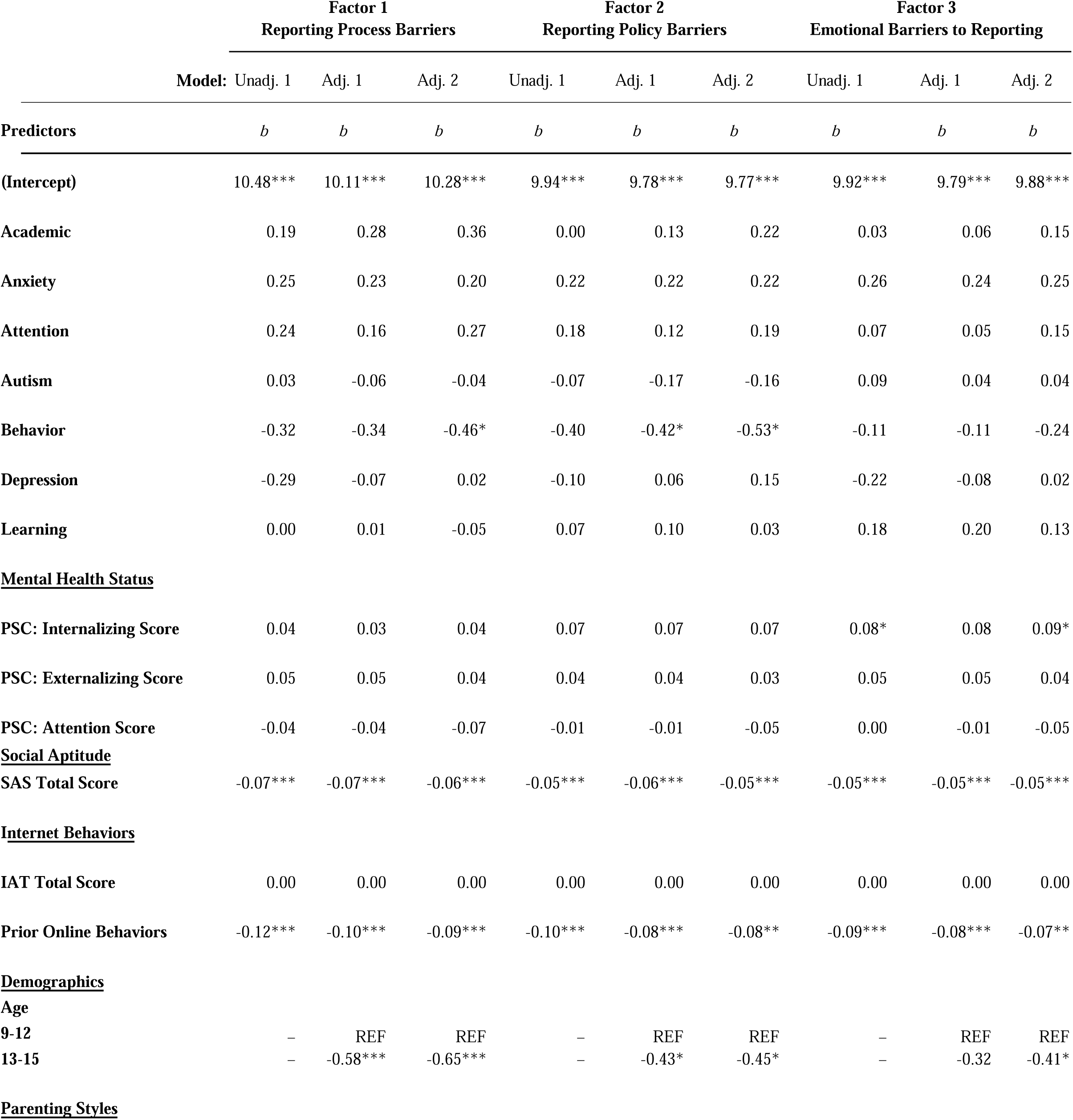
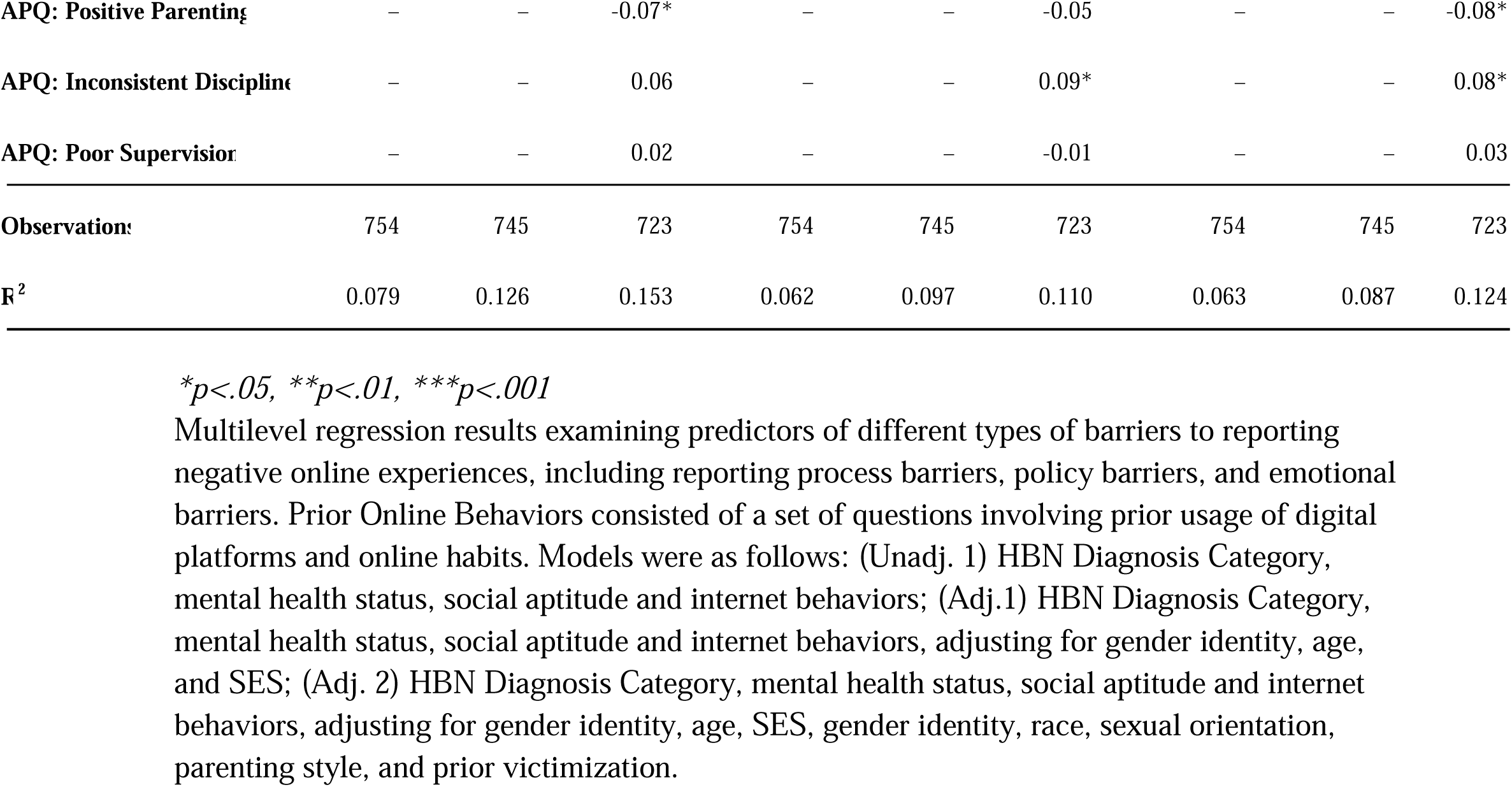
Multivariate regressions of three reporting barriers.

### Mental models and decision-making processes

Qualitative analyses revealed that participants use three components to assess NOE when deciding whether to make a report: (1) degree of malice to the victim; (2) indicators of the perpetrator’s intent; (3) number and severity of the harassment form(s) present. Increased ambiguity in any of these components resulted in elevated levels of uncertainty in the participants when deciding whether to report the presented scenario.

Harassment severity scores were calculated for each hypothetical scenario, described in eTable 3, based on the number and severity of the harassment form(s) present. Higher scores were significantly correlated with increased decision to report (rs=0.308, *p*<.05). There was a slight preference for reporting to an adult (71.74% [20.03%]) over a platform (62.42% [24.31%]). While younger participants (9-12 years; 57.57% [30.97%]) tended to report more often than older participants (13-15 years; 51.99% [31.68%]), they also expressed greater uncertainties about reporting.

### Desired features to create safer online spaces

Less than half (47 [47.00%]) were comfortable reporting NOE on a platform that they frequently use. Increased comfort levels were associated with greater ease in locating the reporting button (66 [66.0%]) and if platforms provided more support to users during (60 [60.0%]) and after (54 [54.0%]) report investigations. Participants desired that platforms provide more information on how users can protect themselves better online (794 [78.69%]). They also expressed a desire for learning more about how to use the platform’s features, such as blocking (746 [76.35%]) and reporting (722 [73.59%]), in general, and through tutorial videos (605 [61.73%]). (eFigure 2-3)

## Discussion

This study examined negative online experiences (NOE) and reporting behaviors in children and adolescents with mental health and neurodevelopmental conditions to provide a deeper understanding into their experiences with NOE. Over one in four participants experienced NOE in the past year, with 68.7% reporting multiple incidents. Aligning with previous research^19,37,38^, inconsistent parenting styles and elevated mental health symptoms increased their likelihood of experiencing NOE. Despite its high prevalence, reporting rates were modest and greatly impacted by three key barriers: Reporting Process Barriers; Reporting Policy Barriers; and Emotional Barriers. Insights from the qualitative follow-up study revealed that younger participants chose to report more frequently than older participants but also experienced more uncertainty in deciding to report. Participants’ decision-making process was heavily based on the degree of malice caused to the victim, perceptions about the perpetrator’s intent, and number and severity of the harassment form(s) present. Overwhelmingly, participants voiced a desire for greater clarity and guidance from platforms about reporting and how to practice greater online safety behaviors.

Appropriate recognition of the different reporting barriers that emerge across developmental stages is crucial for promoting greater online safety. Younger participants (9-12 years) were less likely to report NOE due to being hindered by a lack of understanding about reporting processes and policies. While it is not uncommon for an online user to be in this age group^7,8,26,39–42^, most digital platforms implement minimum age restrictions of 13 to comply with privacy laws (e.g., Children’s Online Privacy Protection Act^43^). As a result, many platforms’ technical, social, and linguistic components may be too complex or mature for younger users to understand^44–46^. These two barriers were also significant obstacles for youths with lower social aptitude, regardless of age, suggesting that confusion and hesitation about reporting may be associated with one’s ability to decode social cues, manage emotions, and understand platform language. Regardless of how challenging it may be to implement age verification processes with full accuracy, there seems to be significant value for platforms to strengthen these processes to protect their users. Our findings support prior work^47^ that suggest that it may be beneficial for platforms to increase clarity and accessibility around the reporting process. Enhancing the visibility and usability of reporting tools can reduce confusion^48^, especially for younger users and those with lower social aptitude. Educational resources, like interactive tutorials and user-friendly guides, may further demystify the process and policies^49^, encourage reporting, and foster safer online spaces for at-risk youth.

Emotional responses such as embarrassment, fear, and worry are commonly reported among individuals who experience NOE^26,37^; however, these reactions appear to disproportionately serve as a reporting barrier for youth with internalizing symptoms (e.g., anxiety, mood dysregulation). Potential solutions for platforms to reduce emotional barriers could include strengthening anonymity protections to prevent retaliation^50^ and tailoring messaging to be developmentally and emotionally appropriate to suit a variety of users^47^. Additionally, since youth frequently turn to their peers for emotional support and practical advice during moments of distress and uncertainty^51,52^, integrating peer-led reporting guidance and support networks^53^ may help normalize help-seeking and reduce levels of uncertainty and hesitation.

While prior studies link certain parenting styles to increased NOE risk^54,55^, our findings suggest they may also reduce youths’ likelihood of seeking help. Our findings found that parenting styles characterized by higher levels of inconsistent discipline and poor supervision strongly predicted increased likelihood of NOE, suggesting that a lack of parental involvement and oversight may leave youth more exposed to online harm. Conversely, children from families with more positive parenting behaviors were less likely to experience any of the three reporting barriers. In conjunction with youths’ preference for reporting to adults over the platform, these findings point to parents and families as a possible avenue for interventions addressing NOE. Current efforts to ameliorate NOE and increase reporting that are either native to online platforms, or exist as separate programs, tend to not incorporate parents and families^56^, despite their importance as a safety net for youth to keep themselves safe online. Thus, developing new approaches to incorporate parents and caregivers may be a potentially impactful approach to both reducing youth exposure to NOE and strengthening their support system.

Comparing our quantitative survey to our in-depth qualitative analysis revealed a key insight that youth show clarity and a willingness to report NOE when discussing hypothetical scenarios, but they show significant hesitation when they are personally victimized in real life. This discrepancy may suggest that while youth recognize certain behaviors as unacceptable or “report-worthy” in theory, real-life reporting is hindered by emotional, social, developmental, or other barriers. In moments of personal distress, fears of retaliation, uncertainty about the process, or lack of confidence may prevent them from taking action— even when they recognize that reporting is the “right” response to the situation. Future research should explore this intention–behavior gap in greater depth. Platforms might consider tools that not only educate and help youth learn about acceptable and unacceptable online behavior, but also provide them with real-time emotional and procedural support to bridge the gap between recognizing NOE and taking action.

### Limitations

Limitations of the study include the sample’s predominantly white and high–socioeconomic status composition. Although preliminary studies have noted differences in prevalence and experience with NOE by racial and/or ethnic identities^57,58^, further research is needed to explore how racial and socioeconomic factors influence youths’ experiences with and responses to such incidents. While online harassment prevalence rates in the present study align with prior literature^8^, many participants reported uncertainty when asked if they had encountered online harassment in the past year. When combined with the finding from the qualitative online bulletin board that youths sometimes have a difficult time discerning what constitutes online harassment, it is possible that rates of online harassment are greater than reported here. Additionally, the present study did not require that participants disclose any details related to their online harassment incident besides following events (e.g., reporting), which may have provided greater clarity on the low reporting rates.

## Conclusion

Youth, particularly those with mental health and neurodevelopmental conditions, face multiple barriers to reporting NOE. Prioritizing user-centered design, digital literacy, and anonymity protections, platforms can create safer environments that encourage reporting and support^47,50^. Future efforts must consider the needs of at-risk populations when developing online safety initiatives by considering users’ emotional, intellectual, and developmental levels, to ensure high effectiveness and equity among all users^48,49^.

## Article information

### Author statement

*Concept and design*: Alexander, Lindsay; Kass, Mirelle; Milham, Michael; Nikolaidis, Aki

*Acquisition, analysis, or interpretation of data:* Abdullah, Najuma; Bagheri-Hamaneh, Sara; Connell, Tatum; Kass, Mirelle; Nikolaidis, Aki

*Drafting of the manuscript:* Kass, Mirelle

*Critical revision of the manuscript for important intellectual content:* Alexander, Lindsay; Freund, Michelle; Milham, Michael; Nikolaidis, Aki

*Statistical analysis:* Bagheri-Hamaneh, Sara; Connell, Tatum; Kass, Mirelle; Nikolaidis, Aki *Administrative, technical, or material support:* Alexander, Lindsay; Kass, Mirelle; Nikolaidis, Aki *Supervision:* Milham, Michael; Nikolaidis, Aki

## Supporting information

eFigure 1. Frequency of online behaviors by age group.

eFigure 3. Thorn questions on platform features.

## Data Availability

All data produced in the present study are available upon reasonable request to the authors.

## Acknowledgements

This research was funded by the Google Trust and Safety Division. This division aims to understand adolescent’s online experiences and their knowledge and trust of reporting mechanisms so that they can create safer products for users. Google provided feedback but did not design, collect, or analyze data, nor did they participate in the writing of the current manuscript. We would like to acknowledge and thank Cascade Strategies for use of their platform to administer the qualitative online bulletin board and for assisting with preliminary analyses conducted on the qualitative data. Cascade Strategies did not design the content nor participate in the writing of the current manuscript. Lastly, we would like to thank the entire study staff for their efforts and dedication to this projectaper, including but not limited to, Katarina Hajder, Najé James, Francesca Fernandez, Kathleen Moskowitz, Carolyn Chadwick, and Ramon Diah. The views and opinions expressed in this article are those of the authors and should not be construed to represent the views of any of the sponsoring organizations, agencies, or U.S. Government.

