## Supplemental materials. for "Negative online experiences and reporting rates in youth with mental health conditions"

### Supplemental methods

**<u>Sample characteristics</u>**

**Healthy Brain Network (HBN)**

HBN[31] is a community-based initiative that provides comprehensive mental health and learning evaluations to children and adolescents (aged 5-21) in New York at no-cost (*N*=6,872 in July 2023). Through HBN's multimodal battery of assessments that collect a wide range of data types (e.g., behavioral, cognitive, genetic, and neuroimaging[31]), participants and their families are able to gain crucial insights into the youth's needs. HBN's comprehensive report streamlines access to appropriate supports, interventions and care for youths and their families whilst concurrently allowing for the generation of a large open-source dataset that captures the broad range of heterogeneity and impairment present in youth developmental psychology.

**Qualitative follow-up study**

Cascade Strategies (https://cascadestrategies.com/) hosted the bulletin board under a confidentiality agreement. The 109 participants were grouped by age (9-12 or 13-15) and asked to answer questions for 30 minutes per day for 3 consecutive days. Questions included hypothetical scenarios of different harassment forms (eTable 3), perceptions of NOE and reporting behaviors, and factors that influence comfort levels with reporting. Researchers from Cascade Strategies performed qualitative text analysis and prepared a summary of key points and trends of the data. We conducted in-depth thematic analyses and content analyses using the frameworks outlined by Braun and Clarke (2006)[59] and Hsieh and Shannon (2005)[60], respectively.

**<u>Data collection characteristics</u>**

**Background and demographics**

This section consisted of 61 items aimed to assess participant's background. Of these, fifty-nine questions came from validated questionnaires, including the Pediatric Symptom Checklist (PSC)[61], Social Aptitude Survey (SAS)[62], Alabama Parenting Questionnaire (APQ)[63] and EU Kids Online survey[8] and two were project-developed.

**General online behaviors**

The section comprised 112 questions assessing participants' overall online behaviors, with 96 from validated questionnaires including items from the EU Kids Online survey[8] and the IAT and 16 project-developed. Eight of the project-developed items measured the frequency of engaging in different online activities on a 5-point Likert scale (never [1] to almost constantly [5]).

**Negative online experiences (NOE)**
This section included 103 questions to assess the frequency of, feelings about, and behaviors following online negative experiences. Ninety-nine questions were sourced from the EU Kids Online survey[8] (e.g., "In the PAST YEAR, has anything EVER happened when you were online that bothered and/or upset you?") and 4 questions were project-developed (e.g., "There is stigma associated with online harassment").

**Online reporting behaviors**
This section included 34 questions to examine reporting behaviors. Nineteen items from the Thorn Barriers to Reporting Survey[7] identified factors deterring minors from reporting, 3 questions were sourced from the EU Kids Online survey[8] and 12 project-developed items assessed attitudes (e.g., "I am unsure how to report") on a 5-point Likert scale from (strongly disagree [1] to strongly agree [5]).

**Types of online harassment**

Cyberstalking: Cyberstalking is the repeated use of electronic communications to harass, threaten, or intimidate another person in ways that make them fear for their safety.[64]

Denigration: Denigration is the deliberate spreading of false, derogatory, or malicious rumors online to damage an individual's reputation, with the intent to discredit and cause harm[14].

Flaming/Roasting: Refers to hostile online exchanges characterized by aggressive argumentation in chat rooms, instant messages, or via email, often incorporating provocative elements such as capital letters, images, and symbols to amplify emotional impact[12,65].

Harassment: Harassment involves repeatedly sending offensive, abusive, or cruel messages, which may also be shared publicly in digital forums or chat rooms[12].

Masquerading/Impersonation: Masquerading or impersonation is the act of breaking into someone's account and while posing as them, sends harmful messages to damage their relationships or reputation[66].

Outing/Doxing: The deliberate act of searching for, gathering, and exposing private or identifying information about an individual online without consent, with the intent to facilitate harassment, cause harm, or inflict humiliation[11].

Social Exclusion: The intentional behavior of preventing or excluding others from participating in online groups or digital communities[12].

Sexual Harassment: Sexual cyberbullying encompasses "any sexually aggressive or coercive behaviour facilitated through the use of electronic media" including the non-consensual sharing of intimate content, sexual extortion, and unwanted sexual communications in digital spaces[13].

Trickery: Trickery refers to instances where an individual shares personal or embarrassing information with someone, only to discover that the information was subsequently disseminated to others without their consent[67].

### Supplemental tables and figures

***eTable 1. Healthy Brain Network (HBN) diagnosis categories***

| HBN Diagnosis Category | Included Clinical Diagnoses |
|---|---|
| Academic | WIAT Numerical Operations < 85; WIAT Word Reading < 85 |
| Anxiety | Agoraphobia; Generalized Anxiety Disorder; Selective Mutism; Separation Anxiety; Social Anxiety (Social Phobia); Specific Phobia; Other Specified Anxiety Disorder; Unspecified Anxiety Disorder |
| Attention | Attention-Deficit/Hyperactivity Disorder (Combined Type); Attention-Deficit/Hyperactivity Disorder (Inattentive Type); Attention-Deficit/Hyperactivity Disorder (Hyperactive/Impulsive Type); Other Specified Attention-Deficit/Hyperactivity Disorder |
| Autism | Autism Spectrum Disorder |
| Behavior | Conduct Disorder (Childhood-Onset Type); Intermittent Explosive Disorder; Oppositional Defiant Disorder |
| Depression | Major Depressive Disorder; Persistent Depressive Disorder (Dysthymia); Disruptive Mood Dysregulation Disorder; Other Specified Depressive Disorder |
| Learning | Specific Learning Disorder with Impairment in Mathematics; Specific Learning Disorder with Impairment in Written Expression |

Classification of mental health and neurodevelopmental conditions in the study sample based on clinical evaluations from the HBN dataset, including diagnoses of anxiety, ADHD, autism spectrum disorder, behavioral disorders, depression, and specific learning disorders.

***eTable 2. Survey details***

| Measure | Description | Citation |
| --- | --- | --- |
| Pediatric Symptom Checklist (PSC) | The PSC is a brief 17-item questionnaire to screen for psychosocial and emotional problems in children. Respondents are asked to rate the best choice that describes them on a 3-point Likert scale (never [0] too often [2]). | Murphy, J. M., Bergmann, P., Chiang, C., Sturner, R., Howard, B., Abel, M. R., & Jellinek, M. (2016). The PSC-17: Subscale Scores, Reliability, and Factor Structure in a New National Sample. Pediatrics, 138(3), e20160038. https://doi.org/10.1542/peds.2016-0038 |
| Alabama Parenting Questionnaire (APQ) | APQ-9 contains subscales used to derive measures of positive parenting, inconsistent discipline, and poor supervision. Respondents were asked to rate 9 items on their agreement with 9 statements on a 5-point Likert scale (never [1] to always [5]). | Elgar, F.J., Waschbusch, D.A., Dadds, M.R. et al. Development and Validation of a Short Form of the Alabama Parenting Questionnaire. J Child Fam Stud 16, 243–259 (2007). https://doi.org/10.1007/s10826-006-9082-5 |
| Social Aptitude Survey (SAS) | The SAS is a 10-item self-report scale that assesses an individual's social skills, attitudes, and ability to navigate various social situations compared to other children of the same age. Items are rated from above to below average. | Liddle, Elizabeth B., Martin J. Batty, and Robert Goodman. "The social aptitudes scale: an initial validation." Social psychiatry and psychiatric epidemiology 44.6 (2009): 508-513. |
| Wechsler Individual Achievement Test (WIAT) | The WIAT is a comprehensive assessment tool of achievement skills, learning disability diagnosis, special education placement, and clinical appraisal. | Wechsler, D. (2005). Wechsler Individual Achievement Test 2nd Edition (WIAT II). London: The Psychological Corporation. |
| Barratt Simplified Measure of Social Status (BSMSS) | The BSMSS is a 5-item self-report measure of social status completed by parents and produces a total score based on educational attainment and occupational prestige. | Barratt, W. (2006). The Barratt simplified measure of social status (BSMSS): Measuring SES. Unpublished manuscript. http://socialclassoncampus.blogspot.com/2012/06/barratt-simplified-measure-of-social.html |
| HBN Diagnosis Category | Mental health and learning disorder diagnoses made by clinicians after completing the full HBN evaluation based on the results of the Kiddie Schedule for Affective Disorders and Schizophrenia (K-SADS) interview, based on DSM-IV criteria. A total of 10 Consensus Diagnoses were provided. | Kaufman, J., et al. (1997). Schedule for affective disorders and schizophrenia for school-age children-resent and lifetime version (K-SADS-PL): Initial reliability and validity data. Journal of the American Academy of Child & Adolescent Psychiatry, 36(7), 980-988. |
| EU Kids Online | Sections from the EU Kids Online survey were used to assess participant's online experiences and exposure to harmful content. | Zlamal, R., Machackova, H., Smahel, D., Abramczuk, K., Ólafsson, K., & Staksrud, E. (2020). EU Kids Online 2020: Technical report. EU Kids Online. |

| | | |
|---|---|---|
| | | https://doi.org/10.21953/lse.04dr94matpy7 |
| Thorn Barriers to Reporting Survey | The survey serves to shed light on factors that may deter individuals from reporting, thereby guiding efforts to improve reporting mechanisms and support systems. | Thorn. (2023). Responding to Online Threats: Minors' Perspectives on Disclosing, Reporting, and Blocking in 2021 |
| Internet Addiction Test (IAT) | The IAT is a 12-item self-report assessment tool that evaluates the severity of self-reported compulsive use of the internet, assessing aspects like the extent of internet use interfering with daily life, work, social relationships, and emotional wellbeing. | Faraci, P., Craparo, G., Messina, R., & Severino, S. (2013). Internet Addiction Test (IAT): which is the best factorial solution?. Journal of medical Internet research, 15(10), e225. https://doi.org/10.2196/jmir.2935 |

This table provides a comprehensive breakdown of the survey instruments used in the study, including the number of items, response formats, and sources of validated measures. The survey comprised multiple sections: (1) Background and Demographics, which included validated instruments like the Pediatric Symptom Checklist (PSC), Social Aptitude Survey (SAS), and Alabama Parenting Questionnaire (APQ); (2) General Online Behaviors, assessing internet use habits and preferences through EU Kids Online items and project-developed questions; (3) Negative Online Experiences, evaluating the nature and emotional impact of past online harassment incidents using EU Kids Online and original questions; and (4) Online Reporting Behaviors, measuring willingness to report online harassment using questions from the Thorn Barriers to Reporting Survey and additional custom items assessing reporting attitudes and barriers.

*eTable 3. Qualitative follow-up study: Hypothetical scenarios*

| Scenario | Part 1 | Harassment Type | Part 2 | Harassment Type |
|---|---|---|---|---|
| Scenario 1 | During Jane's soccer game, Jane scored a goal and did a victory dance. Later that day, Jane received a text from a teammate that included a link to a TikTok video that someone had posted of her doing her victory dance. | Dissing | The next day, she noticed that about 15 of her classmates had posted videos of themselves imitating her victory dance and making fun of her dance skills. | Dissing; Flaming; Roasting |
| Scenario 2 | Nathan writes a poem for his crush, Britney, and decides to send it to her through Instagram direct message (DM). Britney screenshots the poem and texts it to all her friends. The next day, Nathan sees that someone created an Instagram account with the username "NathanlovesBritney". He clicks on this account and sees that only 3 posts are screenshots of the poem he wrote. | Trickery | Nathan notices that the account made a 4th post of his phone number with the caption "text Nathan if you want a poem!" | Trickery; Outing or Doxing |
| Scenario 3 | Stephanie, a high school freshman, notices that John, her friend's older brother who is 19 years old, keeps "liking" her TikTok videos. A few weeks later, John sends her a friend request on Snapchat and BeReal. Stephanie then receives a text from John and starts texting him every day. A week after, they start FaceTiming every night. | Outing or Doxing | On Stephanie's birthday a few days later, John posts on Instagram 3 different screenshots that he had taken of her (without her knowing) while they had been on FaceTime. | Outing or Doxing; Cyberstalking |
| Scenario 4 | Kai, who transitioned from female to male, shared with his 10th grade class that he now identifies as transgender male. For the next month, Kai received over 20 text messages with videos and photos of transgender celebrities from phone numbers that he did not recognize. | Harassment | Even though Kai now identifies as male, his male classmates refuse to add him to the "boys only" group chat. | Harassment; Social Exclusion |

| | | | | |
|---|---|---|---|---|
| Scenario 5 | Natalie receives a friend request on Instagram from an account with the username "sophiajones19". Given that she has a classmate with the name "Sophia Jones" and that the profile photo resembles her classmate, she accepts the friend request. The next day at school, she mentions this Instagram account to Sophia and Sophia responds that that account is not her and that her mom does not let her have an Instagram account. | Masquerading | This account ("sophiajones19") sends her a direct message (DM) asking her for naked or nude photos. | Masquerading; Cyberstalking; Sexual Harassment |
| Scenario 6 | Skylar (who is 15 years old) and James (who is 17 years old) had been dating for 2 years when James broke up with Skylar because he was leaving for college. A month later, Skylar received a text message on WhatsApp, a messaging platform, from Tommy, who was one of James' new college friends. In the text message, Tommy said "James showed all of us your naked photos. Send me more or I'll post these ones online." | Outing or Doxing; Sexual Harassment | Tommy started to call Skylar every day asking for naked photos and started posting inappropriate comments on her Instagram photos. | Outing or Doxing; Cyberstalking; Sexual Harassment |

This table presents six hypothetical scenarios consisting of different forms of harassment. Each scenario consists of two parts (e.g., receiving a mean comment, being impersonated, or experiencing unwanted attention) followed by a follow-up escalation (e.g., the harassment becoming more public, repeated, or more severe). These scenarios were used in qualitative online bulletin board discussions to assess how youth perceive online threats, barriers to reporting, and their likelihood of seeking support.

***eFigure 1. Frequency of online behaviors by age group***

This figure illustrates the frequency of different online behaviors among participants aged 9-12 and 13-15 years old. Data are categorized by communication (e.g., texting, social media), entertainment (e.g., watching videos, gaming), and academic use (e.g., research, homework tools).

***eFigure 2. Comfort levels for reporting harassment based on the platform's features.***

This figure presents participant comfort levels with various platform features related to reporting harassment and negative online experiences. Youth participants (ages 9-15) were asked to indicate how comfortable they would feel reporting an incident if specific platform policies and mechanisms were in place. Responses were categorized into five levels: very uncomfortable, a little uncomfortable, neither uncomfortable nor comfortable, a little comfortable, and very comfortable.

***eFigure 3. Thorn questions on platform features.***

This figure visualizes responses to key questions from the Thorn Barriers to Reporting Survey[7], which assessed youth perceptions of online safety tools, reporting mechanisms, and platform enforcement policies. Participants rated the importance of various safety-related features on a scale from not at all important to extremely important.

#### *eTable 4. Single item regressions of reporting barriers.*

| | I am confident in managing these situations myself. | | | It is important to report these situations to help other kids like me | | |
|---|---|---|---|---|---|---|
| **Model:** | Unadj. 1 | Adj. 1 | Adj. 2 | Unadj. 1 | Adj. 1 | Adj. 2 |
| **Predictors** | *b* | *b* | *b* | *b* | *b* | *b* |
| **(Intercept)** | 2.58*** | 2.47*** | 2.50*** | 3.67*** | 3.87*** | 3.72*** |
| **Academic** | -0.08 | -0.03 | -0.07 | 0.09 | 0.13 | 0.12 |
| **Anxiety** | -0.04 | -0.04 | -0.04 | -0.08 | -0.08 | -0.11 |
| **Attention** | -0.05 | -0.03 | -0.01 | -0.09 | -0.10 | -0.13 |
| **Autism** | -0.04 | -0.05 | -0.06 | 0.29* | 0.32* | 0.29* |
| **Behavior** | 0.03 | 0.06 | 0.04 | -0.08 | -0.11 | -0.07 |
| **Depression** | 0.03 | -0.05 | 0.05 | -0.03 | -0.02 | 0.06 |
| **Learning** | -0.14 | -0.15 | -0.13 | -0.02 | 0.00 | 0.02 |
| **<u>Mental Health Status</u>** | | | | | | |
| **PSC: Internalizing Score** | -0.04* | -0.04* | -0.04* | 0.00 | -0.01 | 0.01 |
| **PSC: Externalizing Score** | 0.00 | 0.01 | 0.01 | -0.06** | -0.06** | -0.05* |
| **PSC: Attention Score** | 0.02 | 0.02 | 0.01 | 0.07** | 0.08*** | 0.08*** |
| **<u>Social Aptitude</u>** | | | | | | |
| **SAS Total Score** | 0.03*** | 0.03*** | 0.03*** | 0.00 | 0.00 | 0.00 |
| **<u>Internet Behaviors</u>** | | | | | | |
| **IAT Total Score** | 0.00* | 0.00 | 0.00 | 0.00 | 0.00 | 0.00 |
| **Prior Online Behaviors** | 0.05* | 0.04* | 0.04* | -0.01 | -0.01 | -0.02 |
| **<u>Demographics</u>** | | | | | | |
| **Age** | | | | | | |
| **9-12** | – | REF | REF | – | REF | REF |
| **13-15** | – | 0.18** | 0.14* | – | -0.09 | -0.01 |
| **<u>Parenting Styles</u>** | | | | | | |
| **APQ: Positive Parenting** | – | – | 0.00 | – | – | 0.04* |
| **APQ: Inconsistent Discipline** | – | – | -0.01 | – | – | 0.00 |
| **APQ: Poor Supervision** | – | – | 0.03 | – | – | -0.05* |
| **Observations** | 787 | 777 | 753 | 776 | 766 | 743 |
| **$R^2$** | 0.103 | 0.135 | 0.142 | 0.031 | 0.056 | 0.081 |

**p<.05, **p<.01, ***p<.001*

This table presents multilevel regression analyses assessing the relationships between individual psychological traits (e.g., depression, anxiety, behavioral tendencies) and two specific self-reported barriers to reporting ("I am confident in managing these situations myself." and "It is important to report these situations to help other kids like me"). Models were as follows: (Unadj. 1) Diagnoses, mental health status, social aptitude and internet behaviors; (Adj.1) Diagnoses, mental health status, social aptitude and internet behaviors, adjusting for gender identity, age, and SES; (Adj. 2) Diagnoses, mental health status, social aptitude and internet behaviors, adjusting for gender identity, age, SES, gender identity, race, sexual orientation, parenting style, and prior victimization. Prior Online Behaviors consisted of a set of questions involving prior usage of digital platforms and online habits.
