## Supplementary figures and images for "Negative online experiences and reporting rates in youth with mental health conditions"

### eFigure 1. Frequency of online behaviors by age group.

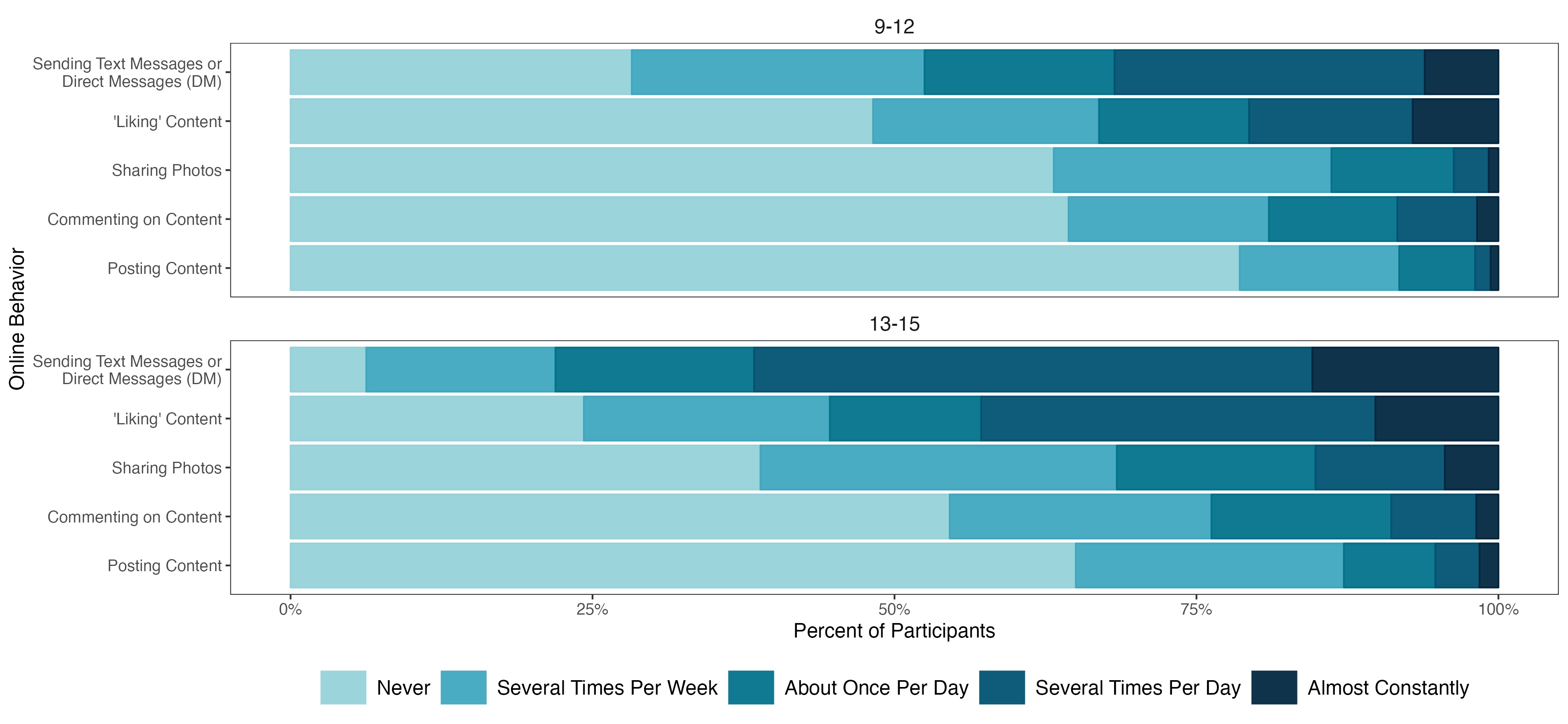

### eFigure 3. Thorn questions on platform features.

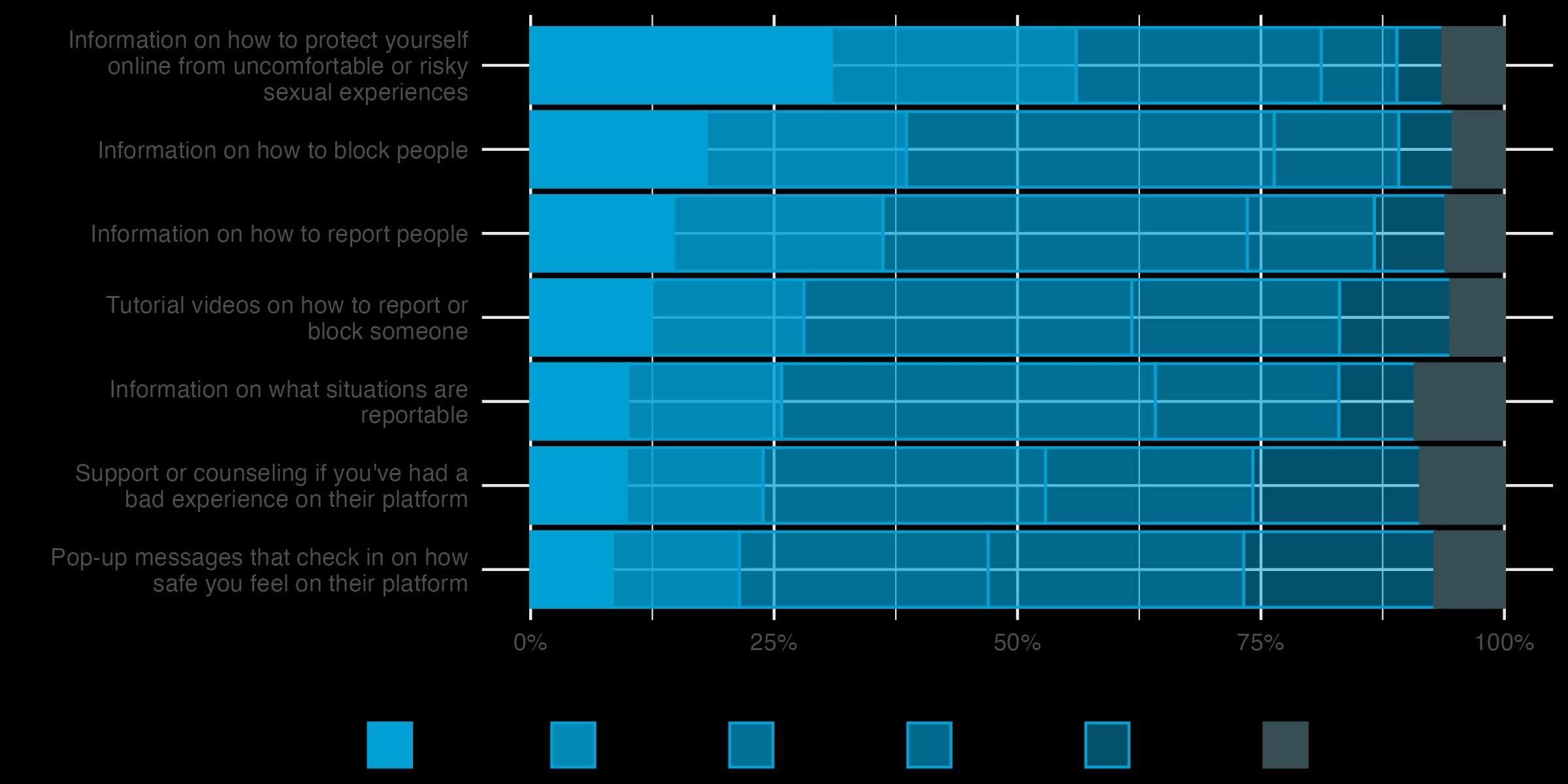

### Supplemental Data 1

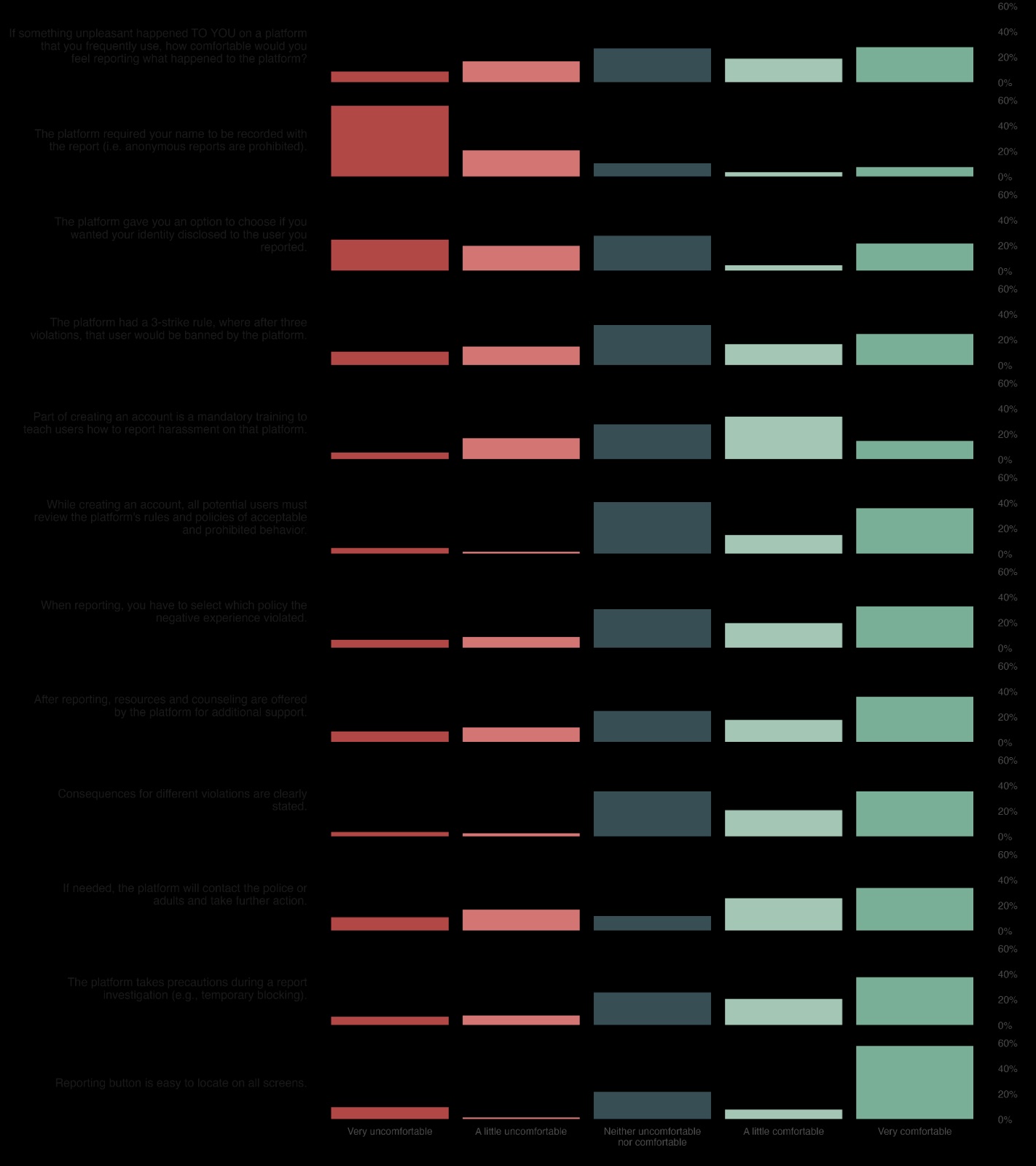
